# Genetic diversity in the Plasmodium falciparum next-generation blood stage vaccine candidate antigen PfCyRPA in Senegal

**DOI:** 10.1101/2024.10.13.24305808

**Authors:** Aboubacar Ba, Laty Gaye Thiam, Mariama Nicole Pouye, Yicheng Guo, Saurabh D. Patel, Seynabou Diouf Sene, Fatoumata Diallo, Rebecca Li, Awa Cisse, Noemi Guerra, Safia Laqqa, Khadidjatou Mangou, Adam J. Moore, Bacary Djilocalisse Sadio, Jean Louis Abdourahim Ndiaye, Alassane Mbengue, Zizhang Sheng, Lawrence Shapiro, Amy K. Bei

## Abstract

The *Plasmodium falciparum* cysteine-rich protective antigen (PfCyRPA) is a promising target as a next-generation blood-stage malaria vaccine and together with PCRCR complex members, the reticulocyte binding-like homologous protein 5 (PfRh5) and the Rh5-interacting protein (PfRipr), are currently being evaluated in clinical trials. PfCyRPA is essential for merozoite invasion and appears to be highly conserved within the *P. falciparum* parasite populations. Here, we used a targeted deep amplicon next-generation sequencing approach to assess the breadth of PfCyRPA genetic diversity in 95 *P. falciparum* clinical isolates from Kédougou, an area with a high seasonal malaria transmission in Senegal. Our data show the dominant prevalence of PfCyRPA wild type reference allele, while we also identify a total of 15 single nucleotide polymorphisms (SNPs). Of these, only five have previously been reported, while the majority of the SNPs were present as singletons within our sampled population. Moreover, the variant read frequency of the identified SNPs varied from 2.6 to 100%, while the majority of the SNPs were present at frequencies greater than 25% in polygenomic samples. We also applied a structure-based modelling approach to thread these SNPs onto PfCyRPA crystal structures and showed that these polymorphisms have different predicted functional impacts on the interactions with binding partner PfRH5 or neutralizing antibodies. Our prediction revealed that the majority of these SNPs have minor effects on PfCyRPA antibodies, while others alter its structure, stability, or interaction with PfRH5. Altogether, our present findings reveal conserved PfCyRPA epitopes which will inform downstream investigations on next-generation structure-guided malaria vaccine design.

## Introduction

Malaria is caused by parasites of the genus *Plasmodium spp*. and remains a major cause of morbidity and mortality, especially in Africa which bears the brunt of over 90% of the disease burden. Despite being a preventable and curable disease, malaria remains a global public health burden with an estimated 608,000 deaths and an associated mortality rate of 14.3 deaths per 100,000 population at risk in 2022. Combined efforts in both preventive and therapeutic measures have significantly reduced the malaria burden over the last two decades. However, this fragile progress has reversed in recent years with the emergence and spread of both insecticide-resistant mosquitos and the antimalarial resistant parasites [1]. This emphasizes the urgent need to accelerate the development of highly effective vaccines against the human malaria parasites [2] which will further support current control measures to reduce the incidence of this disease in endemic countries and strive towards malaria elimination. Malaria vaccine development strategies have recently achieved a milestone following the WHO’s recommendation of the RTS, S/AS01 and R21/Matrix-M (R21) malaria vaccines for the prevention of *P. falciparum* malaria in children living in regions with moderate to high transmission [1, 3, 4]. Both RTS,S and R21 target the circumsporozoïte protein of the *P. falciparum*’s liver stage and have extensively been evaluated in clinical trials and pilot implementation for RTS,S. Primary analysis of R21 phase 3 clinical trial data showed protection of 67-75% against multiple clinical malaria episodes after a 12-month follow-up of fully vaccinated children (5-36 months) [4], while that of RTS, S vaccine is limited by its modest efficacy, as demonstrated in the large phase 3 clinical trial across eight African countries where efficacy was 55.8% in children aged 5–17 months was observed over first year, and waned to 18.3% and 28.2% in infants (6-12 weeks) and children (5-17 months), respectively over 48 months of follow-up [5, 6]. There is an opportunity to complement these first-generation with next-generation vaccines, preferably targeting other stages of the parasite’s life cycle, to complement the existing malaria vaccine toolbox. Such vaccines need to consider genetic diversity at the very early stage of development. Malaria vaccine development has tremendously benefited from the publication of the genome of the *Plasmodium falciparum* [7], which has paved the way for malaria reverse vaccinology [8]. This approach enabled the prioritization of current lead blood-stage malaria vaccine *P. falciparum* reticulocyte binding homolog 5 (PfRh5), the terminal member of the PCRCR complex [9] that binds to erythrocyte receptor Basigin [10]. In addition to PfRh5, members of this complex include the Rh5-interacting protein (PfRipr), Cysteine-rich protective antigen (PfCyRPA), the Plasmodium thrombospondin-related apical merozoite protein (PfPTRAMP), and the cysteine-rich small secreted protein (PfCSS) [9, 11]. Of these, PfRh5 remains the most advanced antigen of the complex in clinical development, having recently completed Phase 2b clinical trials in Burkina Faso, while PfCyRPA and PfRipr are currently being assessed in phase 1 clinical trials (NCT0538547) [12]. Our present study evaluates the breadth of genetic diversity of PfCyRPA and uses structural insights to predict the functional impact of such diversity, contributing to structure-guided vaccine development.

## Results

### Characteristics of study participants

This study was conducted in Kedougou, a Southeastern region of Senegal, with a seasonal malaria transmission from May to November. The study protocol was approved by National Ethics Committee of Senegal (CNERS) (SEN19/36 and SEN23/09), the regulatory board of the Senegalese Ministry of Health and the Institutional Review Board of the Yale School of Public Health (2000025417). Informed consent was obtained from study participants and/or their legal guardians. A total of 94 patients presenting confirmed cases of symptomatic *P. falciparum* infection and recruited in 2019 and 2022 from five healthcare centres in Kédougou, Bandafassi (N = 21), Camp militaire (N = 23), Dalaba (N = 33), Mako (N = 13) and Tomboronkoto (N = 4). Table 1 summarizes the demographic and parasitological characteristics of the study participants. Participants enrolled for this study were aged 2 to 67 years (Median 21.75; SD = 13.11), and there were 59 males and 35 females. We observed significantly different sex ratios, with an overall sex ratio of 1.68 in favour of the males. While no significant difference was observed in the median age across sampling sites, we observed a lower proportion of children (≤10 years) across sites. Moreover, we observed an overall complexity of infection (COI) of 3.65 our study population (**Table 1**).

**Table 1:**
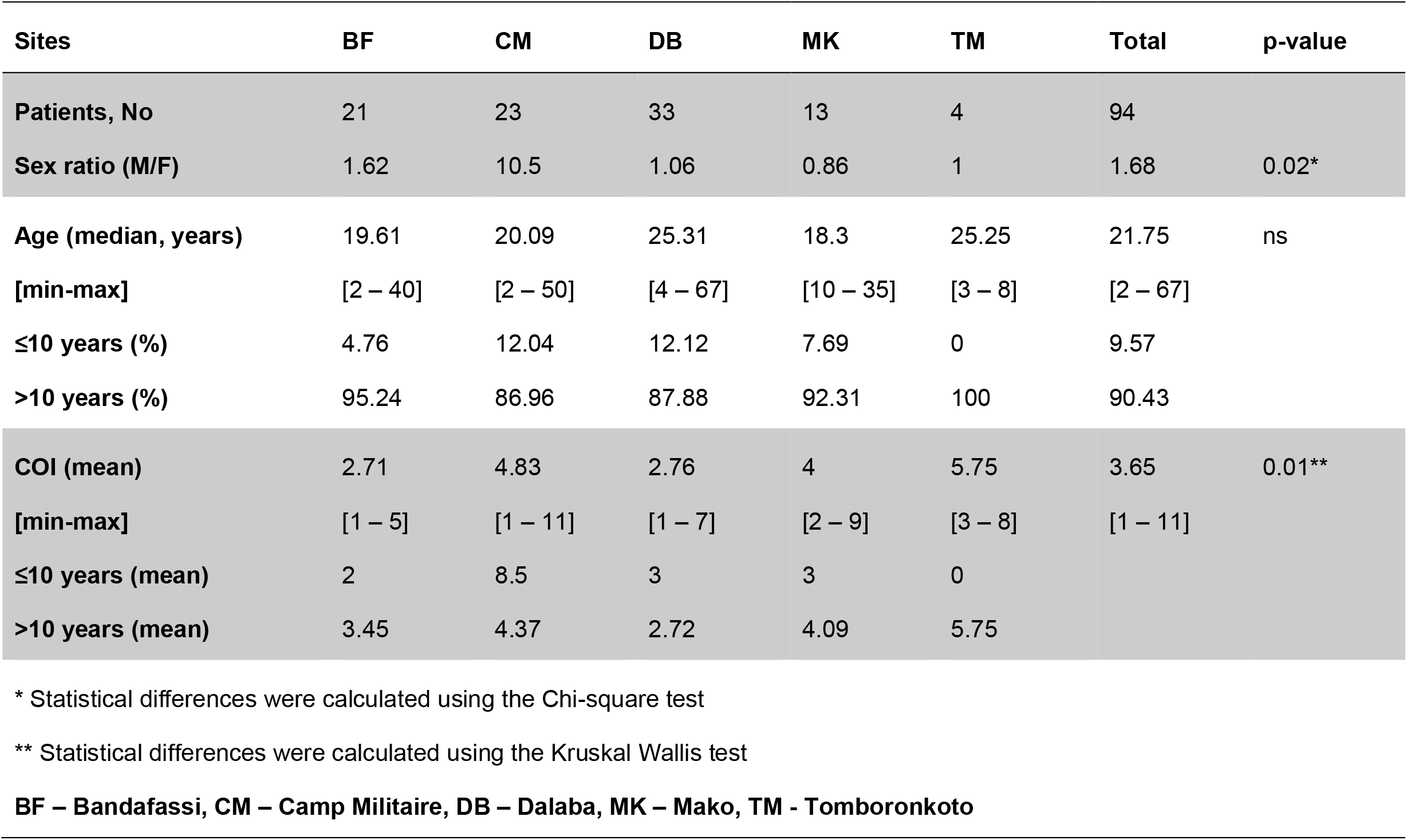
Socio-demographic and parasitological characteristics of study participants.

| Sites | BF | CM | DB | MK | TM | Total | p-value |
| --- | --- | --- | --- | --- | --- | --- | --- |
| **Patients, No** | 21 | 23 | 33 | 13 | 4 | 94 |  |
| **Sex ratio (M/F)** | 1.62 | 10.5 | 1.06 | 0.86 | 1 | 1.68 | 0.02* |
| **Age (median, years)** | 19.61 | 20.09 | 25.31 | 18.3 | 25.25 | 21.75 | ns |
| **[min-max]** | [2 – 40] | [2 – 50] | [4 – 67] | [10 – 35] | [3 – 8] | [2 – 67] |  |
| **≤10 years (%)** | 4.76 | 12.04 | 12.12 | 7.69 | 0 | 9.57 |  |
| **>10 years (%)** | 95.24 | 86.96 | 87.88 | 92.31 | 100 | 90.43 |  |
| **COI (mean)** | 2.71 | 4.83 | 2.76 | 4 | 5.75 | 3.65 | 0.01** |
| **[min-max]** | [1 – 5] | [1 – 11] | [1 – 7] | [2 – 9] | [3 – 8] | [1 – 11] |  |
| **≤10 years (mean)** | 2 | 8.5 | 3 | 3 | 0 |  |  |
| **>10 years (mean)** | 3.45 | 4.37 | 2.72 | 4.09 | 5.75 |  |  |

\* Statistical differences were calculated using the Chi-square test
\*\* Statistical differences were calculated using the Kruskal Wallis test
**BF – Bandafassi, CM – Camp Militaire, DB – Dalaba, MK – Mako, TM - Tomboronkoto**

### Prevalence of SNPs

To determine the degree of PfCyRPA-associated genetic diversity within the population, we employed targeted deep amplicon sequencing using Illumina short-read next-generation sequencing on a NovaSeq6000 platform. Genetic diversity was assessed using a very sensitive threshold (2% variant allele frequency) and applied both qualitative and quantitative metrics to enable accurate SNP discovery and validity. A total of 93 isolates were included in this analysis and the resulting sequences were compared to that of the reference strain (3D7). Overall, 26/93 (28%) of the isolates carried at least one SNP in the PfCyRPA gene relative to the 3D7 reference, which represented the dominant allele 67/93 (72%) within our sampled population (**Fig. 1A**). We identified 15 individual SNPs, of which only five (F41L, V165I, D236N, N270T and V292F) have previously been reported. Additionally, of the novel SNPs reported here, 2 were previously described at the same position but we observed different amino acid substitutions (D236N and N338D). The majority of the novel SNPs were rare and only identified in a single isolate, except for R50C, I196F and K211Q, which were all found in two isolates each. Overall, most of the isolates with a mutant allele carried only a single SNP at a time, while the highest number of SNPs carried by a single isolate (320697) was 4 (R31H; F41L; D236N; V292F). Moreover, the majority of the SNPs reported in this study were rare variants, as only one (V292F) was detected at a prevalence greater than 5%. Of the novel SNPs detected in the study, only two reached a prevalence of at least 2% in the total population I196F (2.2%) and K211Q (2.2%), while the remaining SNPs were all detected at a prevalence of 1.1%, corresponding to a single sample (**Fig. 1A**).

**Figure 1.**
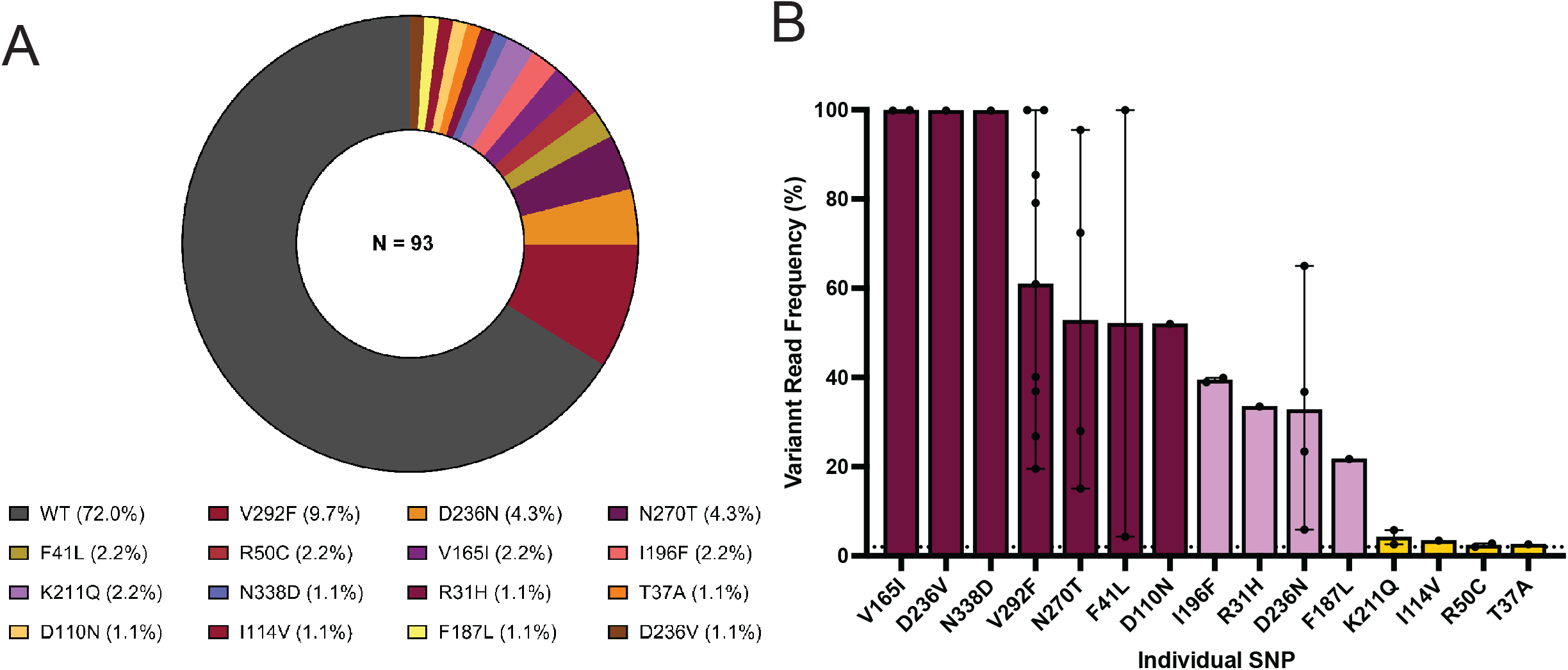
Population prevalence of PfCyRPA SNPs. (A) The prevalence of PfCyRPA-associated SNPs was calculated as the percentage of SNPs detected within the total number of clinical samples in the population (N=93) using a variant allele frequency (VAF) threshold of 2%. PfCyRPA sequencing was performed from pfcyrpa amplicons using the Illumina NovaSeq 6000 sequencing platform and variant analysis was performed using the Geneious Prime software version 23.1.1. The graphs were plotted using the GraphPad Prism version 1.0.2 software. (B) Variant read frequency of PfCyRPA SNPs. Variant read frequency was determined from the sequencing data outputs and calculated as the percentage of the variant reads relative to the coverage at the variant position. The data are presented as bar graphs showing the number of isolates (black dots) with the error bars presenting the minimum and the maximum frequencies for each SNP in complex clinical samples. The SNPs are categorized as low <20% (golden), intermediate 2-25% (lavender) or high frequency SNPs >25% (maroon), based on their respective frequency within the individual complex sample. The dotted line depicts the 2% VAF threshold. The graphs were plotted using the GraphPad Prism version 1.0.2 software.

### Variant frequency of individual SNPs

Given the likelihood of mixed-genotypes infections associated with natural malaria infections in high transmission settings, we measured the complexity of infection (COI) in our sample using *msp1 & 2* genotyping [13]. Overall, only 10 out of the 93 isolates reported here were from monogenomic infections, while the number of polygenomic infections ranges from 1 to 11, with a mean COI of 4 genotypes per isolate. Consequently, we next sought to assess range of variant allele frequencies of these SNPs within these complex infections. This analysis showed that the identified SNPs were distributed at varying frequencies ranging from 2.6 to 100% within the individual isolates; hence the defined classification as low frequency (<5%), intermediate frequency (5–25%) and high frequency (> 25%) SNPs (**Fig. 1B**). Interestingly, 10 out of the 15 SNPs reported here were present at high variant frequencies within the patient sample, while only two SNPs were present at low frequencies. Of these SNPs with high variant frequencies, five were novel (D236V, N338D, D110N, I196F and R31H), while all the previously reported SNPs were present at high frequencies. This increases the confidence that even though rare in the population, these represent SNPs. Two of the novel SNPs (R50C and T37A) were present at low frequencies, while F187L, K211Q and I114V were present at intermediate frequencies (**Fig. 1B**). Out of the 10 isolates with monogenomic infections, only three carried a mutation on *pfcyrpa*, which is present as a single SNP per isolate. Of these SNPs, only I114V (400133) was present at low frequency, while V292F (400116) and N338D (400115) were both present at a high frequency. The majority of the novel SNPs (8/10) described here are present within polygenomic infections.

### Structural modelling of SNPs

We assessed the predicted functional impact of identified polymorphisms by threading the SNPs onto the crystal structure of PfCyRPA, a six-bladed β-propeller protein [14, 15]. These blades, interconnected by loop regions, are each constructed by a four-stranded anti-parallel b-sheet[14][15]. The detailed interactions between PfCyRPA and its binding partners (RH5 and Ripr) or monoclonal antibodies have more recently been reported [11, 16]. Consequently, we used a structure-guided approach to thread the identified SNPs onto the PfCyRPA crystal structure in complex with PfRH5 and earlier characterized mAbs (**Fig. 2A-B**). The threading complex was built by superimposing the structure complexes of PfRH5 bound to its receptor Basigin (PDB id: 4U0Q) or PfCyRPA bound to PfRH5 (PDB id: 6MPV), or mAb Fab fragments PfCyRPA-Cy.003 (PBD:7PI2), PfCyRPA-Cy.004 (PBD:7PHW), PfCyRPA-Cy.007 (PBD:7PHV) and PfCyRPA-8A7 (PBD: 5TIH). The structural threading analysis revealed an even distribution of the identified SNPs between the PfCyRPA internal loops and individual blades (**Table 2**). Of these mutations, four (K211Q, D236N, D236V and N270T) were located within the blade 4. Likewise, blade 6 also harboured four SNPs (R31H, T37A, F41L and N338D) while, three (V165I, F187L and I196F) and two SNPs (D110N and I114V) were respectively located on blade 3 and blade2. However, the blades 1, and 5 each carries a single mutation (**Fig. 2A-B**). Furthermore, our analysis showed four groups of SNPs with different predicted functional outcomes. Interestingly, 7 out of the 15 SNPs reported here are predicted to have minor effect on PfCyRPA structure or binding with PfRH5 or potentially on monoclonal antibody interactions, while another group of SNPs (V165I, I196F, D236N, N270T and V292F) is predicted to partially alter the structure of PfCyRPA. While both D236 and N270 form hydrogen bonds with S233 and N218 residues, respectively, the D236V and N270T mutations alter the PfCyRPA structure flexibility or stability, respectively through steric clashes or interruption of hydrogen bonds. Another group of SNPs, predicted to affect PfCyRPA interaction with PfRH5 and included R50C, F187L and I196F, the former improving binding to PfRH5 by removing the repulsion between R50 and K504 residues (**Table 2**). Intriguingly, 8 mutations may destabilize PfCyRPA (Table 1). One key implication of structure-guided vaccine design relative to the next-generation blood-stage malaria vaccines is to decipher the functional implication of the vaccine candidate-associated genetic diversity to epitope-paratope interactions. We identified a subset of SNPs with a potential impact on inhibitory monoclonal antibody binding, although these interactions are predicted to be very mild and they are not directly in the epitope bound by Cy.003, Cy.004, and Cy.007. Of these SNPs, only three (T37A, F41L, N338D) were mapped to blade 6, which along with blade 2 have been shown to trigger the most inhibitory antibodies [16], while R50C, D110N and I114V bound to loop regions within blade 1 and 2, it is possible that these SNPs impact antibody recognition through structural changes (**Fig. 2C-F**), but such predictions should be functionally validated.

**Table 2.** Predicted functional characteristics of PfCyRPA SNPs. Individual FASTA files of PfCyRPA alleles were threaded through the crystal structure and the impact of the mutant versions of the protein was evaluated for predicted binding affinity of PfCyRPA to its binding partner PfRH5 or neutralizing human mAbs. The positions of the SNPs were determined using structural data from Chen et al., 2017. The binding energy alternation from the SNPs was predicted by FoldX version 5.0. Predicted binding energies are shown for reference and mutant alleles of the protein in Kcal/Mol for each SNP. Changes between the two are shown as ΔΔG (Kcal/mol). A negative ΔΔG indicates a predicted increase in PfCyRPA stability while a positive ΔΔG is associated with a predicted decrease in PfCyRPA stability.

| SNP | Blade | Predicted impact on interaction with RH5 and antibodies Cy.003, Cy.004, and Cy.007 | $\Delta\Delta G$ of CyRPA stability (Kcal/mol) |
| --- | --- | --- | --- |
| R31H | 6 | Minor effect | 1.91 |
| T37A | 6 | Minor effect | 0.45 |
| F41L | 6 | Minor effect | 1.23 |
| R50C | 1 | Minor effect on antibody binding, but may improve RH5 binding by removing repulsion between CyRPA R50 and RH5 K504 | 1.11 |
| D110N | 2 | Minor effect | -2.02 |
| I114V | 2 | Minor effect | 0.57 |
| V165I | 3 | Minor effect; may alter CyRPA structure | -0.99 |
| F187L | 3 | Minor effect on antibody binding, but may affect binding to RH5 | 0.2 |
| I196F | 3 | Minor effect on antibody binding, may affect binding to RH5; may alter CyRPA structure | 9.33 |
| K211Q | 4 | Minor effect | 0.95 |
| D236N | 4 | Add a new N-glycosylation site, minor effect | 0.05 |
| D236V | 4 | Alter local structure through steric clash with S233, minor effect on antibody and RH5 binding | 0.09 |
| N270T | 4 | Minor effect on antibody and RH5 binding; alter CyRPA structure, but abolishes a hydrogen bond between CyRPA N218 and N270 | -0.03 |
| V292F | 5 | Minor effect; may alter CyRPA structure | 8.54 |
| N338D | 6 | Minor effect | 1.75 |

**Figure 2.**
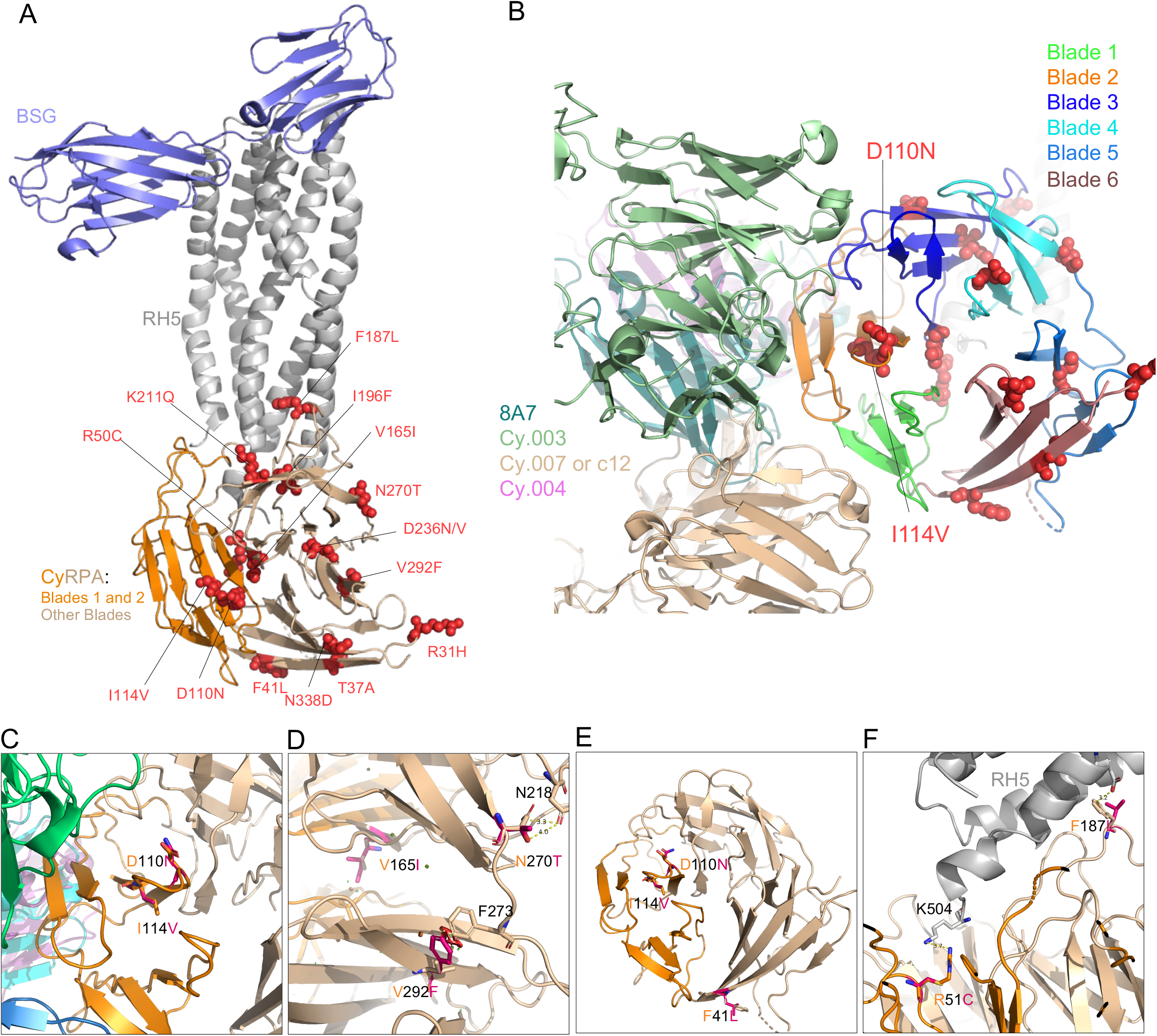
Structure-function predictions for the novel SNPs identified in CyRPA. **(A)** The location of SNPs within the BSG–RH5–CyRPA complex. The complex construction was achieved by superimposing the RH5 of the RH5–BSG complex (PDB ID: 4U0Q) onto the RH5–CyRPA complex (PDB ID: 6MPV). BSG and RH5 are depicted in light blue and grey, respectively. CyRPA is represented in a wheat color, while blade 1 and blade 2 are indicated in dark orange. **(B)** The positions of D110N and I114V relative to monoclonal antibodies (mAbs) with CyRPA blades color coded. Antibodies Cy.003, Cy.004, Cy.007, and 8A7 are represented in green, magenta, blue, and cyan, respectively.(**C**) The positions of D110N and I114V relative to monoclonal antibodies (mAbs). Antibodies Cy.003, Cy.004, Cy.007, and 8A7 are represented in green, beige, magenta, and dark turquoise, respectively. (**D**) Structural modeling revealed that SNPs V165Y, N270T, and V292F influence the conformation of CyRPA. Small red plates signify the potential steric hindrances. (**E**) This panel highlights the SNPs that might confer resistance to antibodies, including F41L, D110N, and I114V. (**F**) SNPs R51C and F187L are shown to directly interact with RH5.

## Discussion

The *P. falciparum* cysteine-rich protective antigen (PfCyRPA) plays a crucial role in merozoite invasion of the human erythrocyte. This antigen has attracted a particular attention as a promising vaccine candidate, as it is essential[17, 18] and accessible to naturally derived human antibodies[19]. Preclinical studies have shown that PfCyRPA induces broadly neutralizing antibody response [15, 16, 20, 21], with a relatively conserved sequence in the various malaria parasites[22, 23], suggesting that a vaccine based on this protein may offer broader protection. However, despite its general conservation, PfCyRPA has some genetic variability, which could limit the effectiveness of a vaccine, as genetic variations in PfCyRPA may allow the parasite to mutate and evade the vaccine-induced immune response. Together with PfRipr, PfCyRPA has recently entered Phase 1 clinical testing (NCT05385471), thus a better understanding of the breadth and functional impact of PfCyRPA-associated polymorphisms in vaccine-induced immune response is needed to prioritize, design and optimize PfCyRPA-based vaccine alleles.

This study was undertaken to assess the extent of PfCyRPA genetic diversity in *P. falciparum* clinical isolates from naturally infected individuals in high malaria transmission settings. Samples reported here were collected from patients diagnosed with *P. falciparum* infections visiting healthcare centres in Kédougou, a Southeastern region of Senegal with high seasonal malaria transmission[24]. A previous study by Ndigwa and colleagues reported an excess of rare variants in proteins within the PfRH5 complex, including PfCyRPA[23]. In this study, the authors used two sequencing strategies, namely capillary Sanger sequencing and whole genome sequencing (WGS), which respectively identified 4 and 10 PfCyRPA-associated SNPs, while only a single SNP was concomitantly discovered by both strategies[23].

Long read sequencing strategies such as Sanger sequencing enable the manual identification of genetic variation and the haplotype calling; however, they are limited by both their overall low throughput and their inability to accurately identify and segregate SNPs in the context of polygenomic infections such as those common in high transmission settings like Kédougou. We previously reported on the high prevalence of polygenomic infections in Kédougou [25]. This trend was confirmed in this current study, with isolates harboring 1 to 11 genotypes, while the mean COI reported here is 4 genotypes per isolate.

To increase our chance of discovering newly emerging and rare PfCyRPA-associated variants, we opted for a targeted deep amplicon sequencing using the Illumina Novaseq 6000 sequencing technology and used a sensitive discovery threshold of 2% for variant calling. We successfully sequenced *pfcyrpa* amplicons from 93 isolates and reported a total of 15 SNPs, of which 10 were novel, while only 5 were reported in previous studies [23, 26]. Interestingly, our current data showed the PfCyRPA reference allele being the most prevalent allele, while the opposite trend was observed for PfRH5[25, 27]. This observation aligns with previous report suggesting a stronger balancing selection pressure on PfRH5 than that on PfCyRPA [19]. Additionally, our findings matched previous reports on the occurrence of an excess of rare variants[23], as the majority of the SNPs reported here were present as singletons (occurring in single isolates), while only three SNPs (V292F, D236N and N270T) were present in more than two isolates. This result emphasizes the power of deep amplicon sequencing strategies in identifying rare genomic variants in polyclonal infections and agrees with previous reports involving the lead blood-stage malaria vaccine candidate, PfRH5[25, 27].

One limitation of the deep amplicon sequencing strategy used here is its inability to resolve individual parasite haplotypes due to the short reads but also to the high complexity of infection in this population. Consequently, for each identified SNP, we assessed the variant read frequency, defined as the percentage of variant reads in the total reads mapped to a given position in the PfCyRPA reference. Given the varying number of genotypes as well as their respective parasitemia in a given isolate, this analysis of the variant reads frequencies enabled us to quantitatively calculate the number of reads with the SNP relative to the total number of reads at its position and therefore classify the SNPs as low (<5%), intermediate (5-25%) and high (>25%) in each given sample. Interestingly, 10 out of the 15 SNPs reported here were present at high frequencies, while 1 and 4 SNPs were respectively present at intermediate and low frequencies. A similar observation was made in our previous study on PfRH5[25], which resulted in a number of SNPs present at low frequencies. While all previously reported SNPs, the most prevalent in the population, were also present at high frequencies, we also showed 4 novel SNPs (R31H, D110N, D236V and N338D) present at high frequencies, each of which were identified as singletons. Of these novel SNPs present at high frequencies, three (R31H, D110N and D236V) occurred in polygenomic infections. Interestingly, the D236V SNP, present as a singleton with a frequency of 99.8% emerged from an isolate with a COI of 4, which emphasizes a hypothesis to further test that the functional implication of this SNP could increase the parasite’s fitness.

Given the relationship between a protein’s structure and its function, we sought to investigate the impact of the identified SNPs in PfCyRPA structure and ultimately predict their functional implication in binding with partner protein PfRH5 or its recognition by neutralizing monoclonal antibodies with known binding epitopes. The crystal structure of PfCyRPA has earlier been solved[14, 15], while more recent studies have solved its structure in complex with neutralizing antibodies as well as binding partners PfRH5 and PfRipr[11, 16]. The functional implication of naturally arising polymorphisms however might be very challenging to investigate within the naturally circulating parasite populations, and mostly in the context of high malaria transmission where individual isolates are often represented as polygenomic infections.

As a primary investigation, we adopted an in-silico approach based on the threading of the observed SNPs onto the crystal structure of PfCyRPA in complex with binding partner PfRH5 or neutralizing antibodies. By superimposing the identified SNPs onto the PfCyRPA structure, we were able to accurately map their distribution and predict their impact on the antigen’s functional structure. Interestingly, in addition to the even distribution of the SNPs between the antigen’s internal loops and blades, there was at least one SNP present in each given blade. Moreover, out of the 15 SNPs located within PfCyRPA blades, 7 were located within blades 3 and 4, which together form much of the interface between PfCyRPA-PfRH5 interaction[11]. As these SNPs were predicted to have either a minor effect on the antigen’s structure or to impact its binding with PfRH5, their functional impact remains a mystery to be solved considering that their location might not be readily accessible to neutralizing antibodies but also given that antibodies occurring in the binding interface between the two antigens are not inhibitory[16, 28]. On the other hand, while a previous study reported the predominance of conformational neutralizing epitopes within the PfCyRPA structure [21], recent data have shown the most of the inhibitory antibody binding epitopes to be located within the blades 1 and 2 of the PfCyRPA structure[16]. Our structural threading analysis showed R50C to be located within blade 1, while both D110N and V165I were located within the blade 2 of PfCyRPA, with both predicted to have a minor effect on PfCyRPA structure.. Moreover, 4 out of the 15 SNPs (R50C, F187L, I196F, N270T) reported here were predicted to have a minor effect on antibody binding to PfCyRPA. However, even if there seems to be no SNPs found within the most critical epitopes of the PfCyRPA so far reported, this should not prevent further investigation on the potential impact of these SNPs, as even though predicted with a minor effect, they could have an important contribution in the parasite overall fitness. Finally, our predictions also reported on the presence of SNPs that can impact the antigen’s functionality in other ways, such as removing repulsive interactions (R50C), disrupting hydrogen bonds (N270T) or causing steric clashes (D236V), while another subset of SNPs has the propensity of either altering (N338D) of introducing (D236N) new glycosylation sites. Given the importance of their structural changes and their key roles in protein stability, folding and solubility, these findings warrant further investigation in order to confirm the functional impact of these SNPs in the context of malaria vaccinology as it relates to PfCyRPA.

Despite the relevance of the reported data from this study relative to the importance of PfCyRPA as a malaria vaccine candidate, there is still room for improvement as it relates to the strategy herein described. The cross-sectional approach described here provides a snapshot of the *pfcyrpa* genetic diversity within the circulating parasite populations. The passive recruitment strategy adopted here could prompt the tendency to only focus on isolates driving the more prominent clinical disease that influences the patient to seek care while overlooking the true natural genetic diversity in isolates from the larger community. As we highlighted in our previous study, future work can address this by sampling across the clinical presentation spectrum, including active surveillance of asymptomatic cases[25]. Another limitation of this study is the notable differences in the number of samples from the individual sites, which also did not enable a site by site comparison of the *pfcyrpa* genetic diversity across the sampling sites. This study was not powered to reflect a thorough assessment of genetic diversity stratified by site, although this would be interesting to explore in depth as there could be differences in the parasite populations circulating within each site due to unique epidemiological characteristics. This could be as a result of their geographical location related to the neighbouring countries with whom the region shares borders (Mali and Guinea Conakry) due to the specific activities of each site (mining and trading activities). Therefore, a larger and more thorough sampling across the entire region could strengthen these preliminary data being reported here. Furthermore, while our sequencing approach has advantages for deep coverage and detection or rare SNPs, it also has its own limitations as the generated short reads, combined with the high complexity of infection in this population, make it difficult to accurately resolve individual parasite haplotypes. Finally, although our structural modelling can imply potential functional impact of the reported SNPs, more functional studies are needed to accurately decipher, if any, the true mechanistic implication of these polymorphisms into the parasites’ fitness and survival. Given the working hypotheses generated from our investigations, this study, will inform downstream biochemical and functional genetic approaches to evaluate the role of each SNP in PfCyRPA function and survival strategies.

## Data Availability

Sequencing Reads associated with this study have been deposited in the NCBI SRA with the BioProject Accession: PRJNA1109877.

## Author Contributions

A.K.B. conceived the experiments. A.K.B., L.S., and Z.S. supervised the research. A.B., L.G.T., M.N.P., S.D.S., F.D., R.L., A.C., N.G., K.M., A.J.M, A.T., B.D.S., and A.M. collected the samples. A.K.B and A.B. assisted with geolocation. A.B., L.G.T., M.N.P. S.D.S., and F.D. conducted the experiments. Y.G., Z.S., S.D.P. performed structure modelling. A.B., L.G.T., S.L. N.G., and A.K.B. analysed the results. A.B., L.G.T., A.K.B. wrote the manuscript. A.B., L.G.T., A.K.B. J.L.A.N., Z.S., S.D.P. and L.S. reviewed and edit the manuscript. All authors reviewed and approved the final manuscript.

## Acknowledgements

We would like to thank Souleymane Ngom from Dalaba, Lt. Dr Charles Latyr Diagne and Lamine Kane from Camp Militaire, Moctar Mansaly and Gerald Keita from Bandafassi, Safietou Sane and Astou Ndiaye from Mako, Adama Gueye from Bantaco, Der Ciss from Tomboronkoto and all the healthcare workers at these sites for their partnership with Institut Pasteur Dakar. We would also like to thank the people of Kédougou for their invaluable contributions to this work. We would like to thank the Yale Center for Genome Analysis (YCGA). We would like Dr Ines VIGAN-WOMAS for her help and assistance in Immunophysiopathology and Infectious diseases Unit at Institut Pasteur Dakar.

## Methods

### Study Sites and sample collection

This study was conducted in Kedougou, a Southeastern region of Senegal, with a seasonal malaria transmission from May to November. Informed consents were obtained from the study participants or their legal guardians and samples were collected following the approved ethical protocol by the National Ethics Committee of Senegal (CNERS) (SEN19/36), the regulatory board of the Senegalese Ministry of Health and the Institutional Review Board of the Yale School of Public Health (2000025417). Samples used in this study were collected through passive case detection from patients visiting healthcare facilities Bandafassi, Bantaco, Camp militaire, Dalaba, Mako, and Tomboronkoto in 2019 and 2022, during the peak of the malaria transmission season (July and August) with malaria-like symptoms. If participants met the enrollment criteria of fever in the past 24 h, an axillary temperature ≥38°C and/or a positive *P. falciparum* malaria diagnosis from a rapid diagnostic test (RDT) and microscopy, they were offered the opportunity to enroll in the study. After informed consent was obtained, a venous blood sample was drawn into EDTA vacutainers and samples were transported at room temperature to the laboratory for processing; no more than 6 hours between draw and processing.

### DNA extraction, PCR amplification and NGS Library & Sequencing

DNA was extracted from infected erythrocyte pellets using the ZYMO Quick-DNA Miniprep Kit (D3024) following the manufacturer’s instructions. The extracted DNA samples were eluted in 30ul of nuclease free water and stored at -20°C prior to PCR amplification. For PCR amplification, *PfCyRPA*-specific primers were designed using the Geneious Prime software version 23.1.1. The *PfCyRPA* 3D7 reference sequence (PF3D7_0423800, PlasmoDB [29]) was used as template for primer designing and the amplification was performed using a classic PCR protocol. The PCR was done by using Phusion® High-Fidelity DNA Polymerase (Catalog: M0530L, 50X higher fidelity than Taq). **Supplemental table 1** shows the primer pairs and the **Supplemental Table 2 and 3** the PCR conditions and PCR program respectively used for *PfCyRPA* amplification. Following successful amplification, *PfCyRPA* sequences were bead-purified (Omega) and quantified using a Qubit 2.0 fluorometer; and subsequently adjusted to equivalent concentration. Sequencing library preparation was performed with the Nextera XT using unique dual indexes (UDIs) and subjected to a subsequent bead-purification. DNA libraries were quantified by qPCR using Roche KAPA Library Quantification Kit. All samples were normalized to a final concentration of 4 nM. The 96 samples quantified and normalized were pooled into 8-sub-pools, which were further bead-purified and quantified using a KAPA qPCR. The 8 sub-pools were further normalized and combined in equal quantities to form one final pool. This final pool was sent to the Yale Center for Genome Analysis (YCGA) for sequencing on an Illumina NovaSeq 6000 platform with targeted coverage of 500,000 reads per sample.

### Data processing and polymorphism analysis

De-multiplexed forward and reverse sequencing reads obtained for each sample were individually imported to Geneious Prime and paired sequences were obtained using the Illumina paired end setting. Paired sequences were subsequently trimmed using BBDuk plugin. A minimum quality score (Q) of 30 was set for the trimming with a minimum length of 75 base pairs, as we were expecting reads around 150 base pairs. Trimmed sequences were aligned with the 3D7 reference sequence that had been annotated with all known non-synonymous mutations. Single nucleotide polymorphisms (SNPs) annotation was performed using five iterations the criteria for SNP calling was set to a minimum frequency of 0.02 (2%) and 1000 read coverage. Sequence data and SNP analysis was performed by at least 3 individuals for each sample.

### Structural modelling of PfCyRPA-associated SNPs

The structures of PfCyRPA and PfRH5 were downloaded from the Protein Data Bank (PDB, https://www.rcsb.org/). PfRh5-PfCyRPA complex was constructed using Pymol (PDB ID: 4U0Q and 6MPV) and structural predictions with mAb binding was performed using PDB IDs 5TIH, 7PI2, and 7PHW). Individual FASTA files containing amino acid sequences of PfCyRPA and individual novel SNPs were generated. These amino acid sequence files were threaded onto the crystal structure of PfCyRPA in complex with binding partner PfRH5 and/or known monoclonal antibodies. Pymol version 2.3.2 was used to predict the effect and to plot the structural location of each SNPs. The structural effect of the mutant versions of the protein were evaluated in terms of biochemical properties such as hydrogen bonding patterns, steric interactions, and predicted binding affinity between the mutant version of the protein and the Basigin receptor the binding energy alternation for SNPs were predicted by FoldX.

