## Supplementary Tables for "Genetic diversity in the Plasmodium falciparum next-generation blood stage vaccine candidate antigen PfCyRPA in Senegal"

**Supplementary Table 1: PfCyRPA primers**

| ***Gene*** | ***PCR Round*** | ***Primer*** | ***Lab Oligo #*** | ***Sequence*** |
| --- | --- | --- | --- | --- |
| ***CyRPA***  ***PF3D7_0423800*** | Primary | Forward | oAMB451 | AACATTATGATTATCCCTTTTCATA |
|  | Primary | Reverse | oAMB453 | TGTCTACTCATAGTTAGCATAGTAT |

The primers were designed in Geneious Prime software version 23.1.1 based on the reference gene uploaded from PlasmoDB.

**Supplementary Table 2: PCR Reaction Mix**

| **Components** | **Final Conc. In 25µL** | **Vol. for a 25µL reaction** |
| --- | --- | --- |
| **H_2_O** | -- | 15.25 |
| **5X Phusion HF Buffer** | 1X | 5 |
| **50mM MgCl_2_** | 2.0µM | 1.0 |
| **10mM dNTP Mix** | 0.2mM | 0.5 |
| **Fwd Primer (10**µ**M)** | 0.2µM | 1 |
| **Rev Primer (10**µ**M)** | 0.2µM | 1 |
| **DNA sample** | -- | 1 |
| **Phusion High-Fidelity** | 1.0unit/50µL | 0.25 |

The PfCyRPA is amplified by using the Phusion High-Fidelity DNA Polymerase User’s Guide (Catalog # M0530L).

**Supplementary table 3: PCR program**

| **Step** | **Cycle** | **Temperature (^o^C)** | **Time**  **(Minutes)** | **Number of Cycles** |
| --- | --- | --- | --- | --- |
| **1** | Initial Denaturation | 94 | 5 | 1 |
| **2** | Denaturation | 94 | 0.5 | 30 |
| **3** | Annealing | 58 | 0.5 |  |
| **4** | Extension | 68 | 1.5 |  |
| **5** | Final Extension | 68 | 5 | 1 |
| **6** | Refrigeration | 12 | HOLD | --- |
